# Spatially refined time-varying reproduction numbers of SARS-CoV-2 in Arkansas and Kentucky and their relationship to population size and public health policy, March – November, 2020

**DOI:** 10.1101/2021.05.26.21257862

**Authors:** Maria D. Politis, Xinyi Hua, Chigozie A. Ogwara, Margaret R. Davies, Temitayo M. Adebile, Maya P. Sherman, Xiaolu Zhou, Gerardo Chowell, Anne C. Spaulding, Isaac Chun-Hai Fung

**Affiliations:** Arkansas Center for Birth Defects Research and Prevention, Department of Epidemiology, University of Arkansas for Medical Sciences, Little Rock, Arkansas; Department of Biostatistics, Epidemiology and Environmental Health Sciences, Jiann-Ping Hsu College of Public Health, Georgia Southern University, Statesboro, Georgia; Department of Geography, AddRan College of Liberal Arts, Texas Christian University, Fort Worth, Texas; Department of Population Health Sciences, School of Public Health, Georgia State University, Atlanta, Georgia; Department of Epidemiology, Rollins School of Public Health, Emory University, Atlanta, Georgia

**Keywords:** COVID-19, SARS-CoV-2, Reproduction number, rural, policy

## Abstract

**Purpose:** To examine the time-varying reproduction number, *R_t_*, for COVID-19 in Arkansas and Kentucky and investigate the impact of policies and preventative measures on the variability in *R_t_*.

**Methods:** Arkansas and Kentucky county-level COVID-19 cumulative case count data (March 6-November 7, 2020) were obtained. *R_t_* was estimated using the R package ‘EpiEstim’, by county, region (Delta, non-Delta, Appalachian, non-Appalachian), and policy measures.

**Results:** The *R_t_* was initially high, falling below 1 in May or June depending on the region, before stabilizing around 1 in the later months. The median *R_t_* for Arkansas and Kentucky at the end of the study were 1.15 (95% credible interval [CrI], 1.13, 1.18) and 1.10 (95% CrI, 1.08, 1.12), respectively, and remained above 1 for the non-Appalachian region. *R_t_* decreased when facial coverings were mandated, changing by -10.64% (95% CrI, -10.60%, -10.70%) in Arkansas and -5.93% (95% CrI, -4.31%, -7.65%) in Kentucky. The trends in *R_t_* estimates were mostly associated with the implementation and relaxation of social distancing measures.

**Conclusions:** Arkansas and Kentucky maintained a median *R_t_* above 1 during the entire study period. Changes in *R_t_* estimates allows quantitative estimates of potential impact of policies such as facemask mandate.

## INTRODUCTION

Coronavirus disease 2019 (COVID-19), caused by severe acute respiratory syndrome coronavirus 2 (SARS-COV-2), was first reported in humans in Wuhan in December 2019. From the early stages of the pandemic to November 2020, there has been a rise in both cases and deaths among states that contain large rural areas in the United States (US).^1^ Arkansas, one of eight states that did not implement a stay-at-home order, and Kentucky, a state that has been more proactive from the beginning of the pandemic, are two southern states that have very similar COVID-19 morbidity and mortality rates, yet differed in their approach in addressing this pandemic. Both states have regions that are classified as rural (the Delta in Arkansas and Appalachia in Kentucky), which face higher percentages of health disparities and socioeconomic stress compared to their respective state counterparts.

In both states, disparities in rurality, poverty, health conditions, and healthcare access have a significant role. In Arkansas, 41% of Arkansans live in rural counties,^2^ compared to only 14% of the US population who live in nonmetropolitan counties. In Kentucky, 25.3% of individuals in Appalachia live in poverty compared to 15.3% in non-Appalachia.^3^ These rural communities face challenges with the pandemic and may be unsuited to handle large surges within their healthcare systems.^4, 5^ Fifty percent of rural residents are at a higher risk of hospitalization and serious illness if they became infected with COVID-19 compared to 40% of metropolitan residents because of pre-existing health conditions.^6^ Rural residents are more likely to be older, poorer, and have more comorbidities including obesity, diabetes, hypertension, heart disease, and chronic lower respiratory disease than urbanites.^6–10^

The states of Arkansas and Kentucky were chosen for this study’s time period due to the increasing incidence of COVID-19 in the southern US. We also wanted to highlight two southern states that share similar cultural heritages, but are of different political climates in 2020 (a Republican governor in Arkansas and a Democratic governor in Kentucky). The time-varying reproduction number, *R_t_*, represents a pathogen’s changing transmission potential over time. As the average number of secondary cases per case at a certain time *t*, *R_t_*>1 indicates sustained transmission and <1 epidemic decline.^11–13^ Examining the *R_t_* among these two states will provide a better indication of COVID-19 transmission, especially among vulnerable rural areas. Our study aimed to estimate the *R_t_* for COVID-19 within Arkansas and Kentucky and to compare the *R_t_* among the two states, as well to determine if it differs among the urban and rural areas of each state, and to investigate the impact of policies, and preventative and relaxation measures on the *R_t_*.

## METHODS

### 2.1 Data acquisition

Using data from the New York Times GitHub data repository,^14^ we downloaded the cumulative confirmed case count from March 6 – November 7, 2020, for Arkansas and Kentucky, including the counties located in each state. We used the Delta Regional Authority^15^ and the Appalachian Regional Commission^16^ to classify the counties in Arkansas as Delta and non-Delta, and Appalachian and non-Appalachian in Kentucky. A detailed list of all 75 and 120 counties of Arkansas and Kentucky are provided in **Supplementary Tables 1** and **2**. The first case in Arkansas was reported on March 11, 2020, and the first case in Kentucky was reported on March 6, 2020. The study cutoff point was November 7, 2020. The management of negative incident case counts is described in **Appendix A**. We merged the county-level data to obtain the regional-level data (Delta, non-Delta, Appalachian, and non-Appalachian). To generate *R_t_*, from the reported cumulative case count numbers, we utilized the daily number of new confirmed COVID-19 cases. We accessed 2019 county-level population data for Arkansas and Kentucky from the U.S. Census Bureau.^17^

For sensitivity analysis, statewide hospitalization data for Arkansas and Kentucky, were downloaded from the COVID Tracking Project.^18^ The first date of report was April 1, 2020 for Arkansas and April 10, 2020 for Kentucky. Due to an observed weekend effect, the 3-day moving average was applied to both hospitalization datasets before they were further analyzed.

We downloaded the executive orders from the governors’ offices of each state and identified the date of the implementation and relaxation of public health interventions in each state respectively (**Table 1**).

**Table 1.** COVID-19-Related Policies and Measures Implemented in Arkansas and Kentucky, March – October, 2020.

| **Label in R<sub>t</sub> Policy Plot** | **Date** | **Implemented Policies and Relaxation Measures** |
| --- | --- | --- |
| **Arkansas** |  |  |
|  | March 17 | Schools Closed. |
| A | March 19 | Closed dine-in activities at bars and restaurants, gyms and indoor entertainment venues, and schools until April 17, 2020. |
| B | March 23 | Restricted gatherings to 10 people or fewer. |
| C | April 4 | Required businesses, manufacturers, construction companies, and places of worship to implement social distancing protocols, such as: limiting the number of people who might enter a facility at once, marking off six-foot increments if lines formed, providing hand sanitizer or other disinfectant at or near the entrance, using contactless payment systems if the business engaged in retail or disinfecting all portals and pens, and posting a sign at the entrance informing those who entered that they should maintain a six-foot distance and avoid entering if they had a fever or cough. |
|  | April 30 | Governor Hutchinson announced that gyms and fitness centers can reopen on May 4. |
|  | May 1 | Governor Hutchinson announced that barber, cosmetology, massage therapy, body art, and medical spa services may resume operations on May 6. |
|  | May 5 | Executive Order Regarding the Public Health Emergency Concerning COVID-19, For the Purpose of Renewing the Disaster and Public Health Emergency to Prevent the Spread of and Mitigate the Impact of COVID-19. |
| D | May 11 | Dine-in operations continue for restaurants. |
|  | May 15 | Governor Hutchinson announced that as of May 18, 2020, all businesses, with the exception of bars, will be permitted to open in the state. |
|  | May 18 | Governor Hutchinson announced that bars associated with restaurant facilities may open on May 19, 2020, while freestanding bars not associated with restaurants may open with restrictions on May 26, 2020. |
|  | June 2 | State of Emergency Declared. |
| E | June 10 | Governor Hutchinson announced that the state will be moving into Phase 2 of reopening beginning on June 15, 2020. Under Phase 2, social distancing and facial coverings are still recommended, and restaurants and businesses will be allowed to operate at two-thirds capacity, as opposed to the one-third capacity allowed during Phase 1. |
|  | June 18 | Declared an End to the State of Emergency declared on June 2. |
|  | June 29 | Governor Hutchinson has paused further reopening of Arkansas businesses as the number of coronavirus cases in the state continue to spike. |
|  | July 6 | Cities permitted to implement ordinances requiring face coverings to help curb the spread of COVID-19. Previously, only the Governor could mandate the wearing of face coverings, as cities and counties could not take more restrictive measures than those issued by the state government, per Executive Order 20-37. Arkansas does not have a state-wide face covering mandate. |
| F | July 20 | Required use of face coverings/masks in public. |
|  | August 14 | Executive Order to Renew the Disaster and Public Health Emergency to Mitigate the Spread and Impact of COVID-19. |
| G | August 24 | Schools reopened for in-person instruction. |
|  | October 13 | Executive Order to Renew the Disaster and Public Health Emergency to Mitigate the Spread and Impact of COVID-19. |
| **Kentucky** |  |  |
|  | March 6 | State of Emergency Declared. |
| A | March 16 | Schools Closed.<br>Restaurants ceased in person dining. |
|  | March 23 | Closed all in-person retail businesses that were not lifesustaining.<br>Ceased all elective medical procedures. |
|  | March 26 | Ceased all non-life-sustaining businesses in-person services. |
|  | March 28 | Governor Beshear announced that Kentuckians could still go to Tennessee for work, to take care of a loved one or even buy groceries if it was closer, but asked that unnecessary travel to Tennessee end. |
| B | March 30 | Issued order that restricted out-of-state travel, with four exceptions: 1) traveling to other states for work or groceries, 2) traveling to care for loved ones, 3) traveling to obtain health care and 4) traveling when required by a court order. |
|  | April 2 | Expanded recent order restricting travel to include people from out of state coming into the commonwealth. Anyone from out of state had to follow the same travel restrictions as Kentuckians. If people wanted to stay in Kentucky with a family member or friend for the duration of the COVID-19 crisis, that would be okay, but they needed to quarantine for 14 days when they got here and would not travel anywhere else. |
|  | April 3 | All Kentucky State Parks would no longer be open for overnight stays. |
| C | April 4 | Adopted on a voluntary basis the new guidance from the U.S. Centers for Disease Control and Prevention (CDC) |
|  |  | recommending that people wear cloth masks in some situations. |
|  | April 9 | Ordered Natural Bridge and Cumberland Falls state resort parks to close. |
|  | April 20 | Governor Beshear advised the commonwealth's education leaders to keep facilities closed to in-person instruction for the rest of the school year. |
|  | May 6 | Governor Beshear issued new executive order that continued to ban anyone with a positive or presumptively positive case of COVID-19 from entering Kentucky, except as ordered for medical treatment. It also kept in place requirements of social distancing on public transportation. Those traveling from out of state into Kentucky and staying were being asked to self-quarantine for 14 days. |
|  | May 11 | Everybody working for an essential business that was reopening should be wearing a mask. |
| D | May 14 | Groups of 10 people or fewer could gather. |
| E | July 9 | Required use of face coverings/masks in public. |
|  | July 20 | Cabinet for Health and Family Services issued new order that pulled back on guidance covering social, non-commercial mass gatherings.<br>The Kentucky Department of Public Health issued a new travel advisory that recommended a 14-day self-quarantine for travelers who went to any of eight states – Alabama, Arizona, Florida, Georgia, Idaho, Nevada, South Carolina and Texas – that were reporting a positive coronavirus testing rate equal to or greater than 15%. The advisory also included Mississippi, which was quickly approaching a positive testing rate of 15%, and the U.S. Commonwealth of Puerto Rico. |
|  | July 27 | Announced the closing of bars for two weeks, effective, Tuesday, July 28.<br>Announced that restaurants would be limited to 25% of pre-pandemic capacity indoors; outdoor accommodations remain limited only by the ability to provide proper social distancing. Recommended that public and private schools avoided offering in-person instruction until the third week of August. |
|  | August 6 | Extended the state's mandate requiring face coverings in some situations for another 30 days. |
|  | August 10 | Governor Beshear recommended that schools waited to begin in-person classes until Sept. 28. |
|  | August 11 | Issued an executive order allowing bars and restaurants to operate at 50% of capacity, as long as people could remain six feet from anyone who was not in their household or group. Bars and restaurants would be required to halt food and beverage service by 10 p.m. and close at 11 p.m. local time. |
|  | August 12 | Governor Beshear offered an update on his administration's travel advisory, which recommended a 14-day self-quarantine for Kentuckians who traveled to states and territories that were reporting a positive coronavirus testing rate equal to or greater than 15%. The current areas meeting this threshold included Florida, Nevada, Mississippi, Idaho, South Carolina, Texas, Alabama and Arizona. |
|  | September 4 | Extended the state's mandate requiring face coverings in some situations for another 30 days. |
| F | September 28 | Schools reopened with in-person instruction. |
|  | October 6 | Extended the state's mandate requiring face coverings in some situations for another 30 days. |

### 2.2 Statistical analysis

*R_t_* was estimated using the instantaneous reproduction number method as implemented in the R package ‘EpiEstim’ version 2.2-3. This measure was defined by Cori et al.^11^ as the ratio between *It*, the number of incident cases at the time *t*, and the total infectiousness of all infected individuals at the time *t*. This method has been implemented worldwide in multiple studies to estimate the *R_t_* of SARS-CoV-2 and is briefly described in **Appendix B**.^19–26^ We shifted the time series by nine days backward (assuming a mean incubation period of 6 days and a median delay to testing of 3 days)^27^ for generating *R_t_* by the assumed date of infection,^13^ and we specified the serial interval (mean = 4.60 days; standard deviation = 5.55 days).^28^ Besides using the 7-day sliding window, we also analyze *R_t_* by the different non-overlapping time periods when different combinations of non-pharmaceutical interventions have been implemented, known as policy change *R_t_* (PC*R_t_*) thereafter. We estimated the 1-week sliding window *R_t_* and PC*R_t_* for both states at the state and regional levels. We calculated the median *R_t_* difference percentage changes and the 95% credible interval (CrI), comparing with the previous policy interval, by bootstrapping (1000 random samples for each *R_t_* distribution) for each state-level PC*R_t_*, each respective state region, and the hot-spot analyses for each state (**Supplementary Tables 3-6)**.

We also performed the similar analysis at the county-level in which we identified as hot spots based on the reported data and local news (**Appendix C**). For Arkansas, we analyzed data from Washington, Benton, Lincoln, and Yell Counties, respectively, and combined data from Washington County and adjacent Benton County for analysis as they are one metropolitan area (**Supplementary Figure 1**). For Kentucky, we analyzed Jefferson, Shelby, Elliott, and Warren Counties, respectively, and combined data from Jefferson County and adjacent Shelby County for analysis as they are one metropolitan area (**Supplementary Figure 2**).

A sensitivity analysis was performed to estimate 1-week sliding window *R_t_* utilizing statewide hospitalization data (**Appendix D**).

We conducted linear regression between the log10-transformed per capita cumulative case count and the log10-transformed population size,^29, 30^ at four different dates: May 7^th^, July 7^th^, September 7^th^, and November 7^th^. See **Appendix E** for details and results.

Statistical analysis was performed using R 4.0.3 (R Core Team, R Foundation for Statistical Computing, Vienna, Austria). Maps were created using ArcGIS Pro Version 2.4.0 (Esri, Redlands, CA, USA), with color codes arranged according to quintiles of the values.

### 2.3 Ethics

The Georgia Southern University Institutional Review Board made a non-human subjects determination for this project (H20364) under the G8 exemption category.

## RESULTS

As of November 7, 2020, there were 119,057 cumulative confirmed COVID-19 cases in Arkansas (57,836 for Delta and 61,221 for non-Delta) and 122,024 cases in Kentucky (27,480 for Appalachian and 94,544 for non-Appalachian). **Supplementary Figures 3** and **4** present the spatial variation of cumulative case count and cumulative incidence per 100,000 population by county in Arkansas and Kentucky at four different dates: May 7^th^, July 7^th^, September 7^th^, and November 7^th^, 2020, respectively.

### 3.1 Rt estimates at the state and regional level

Overall, the median *R_t_* for Arkansas and Kentucky at the end of the study were 1.15 (95% CrI, 1.13, 1.18) and 1.10 (95% CrI, 1.08, 1.12), respectively. Between both states, the *R_t_* estimates followed similar patterns. However, they were different when examining certain policy changes.

From March 11 to November 7, 2020, Arkansas revealed two major surges of new cases in July and October (**Figure 1**). The 7-day sliding window *R_t_* estimates in Arkansas was high at the beginning, nearing an *R_t_* estimate of 3, dropping below 1 in mid-April, and having peaks above 1 for a few months before steadily staying around 1. At the end of the study, the median 7-day sliding window *R_t_* estimate was 1.15 (95% CrI, 1.13, 1.18). In the Delta region, the 7-day sliding window *R_t_* estimates had more pronounced decreased peaks in mid-May and mid-June, whereas the non-Delta region had two peaks below 1 in the early stages, an increased peak in mid-May that was above 1, and then stabilized around 1. At the end of the study, the Delta and non-Delta median 7-day sliding window *R_t_* estimates were 1.14 (95% CrI, 1.10, 1.17) and 1.17 (95% CrI, 1.13, 1.20), respectively, with both regions demonstrating extensive community transmission of SARS-CoV-2, with a median *R_t_* >1.

**Figure 1.**
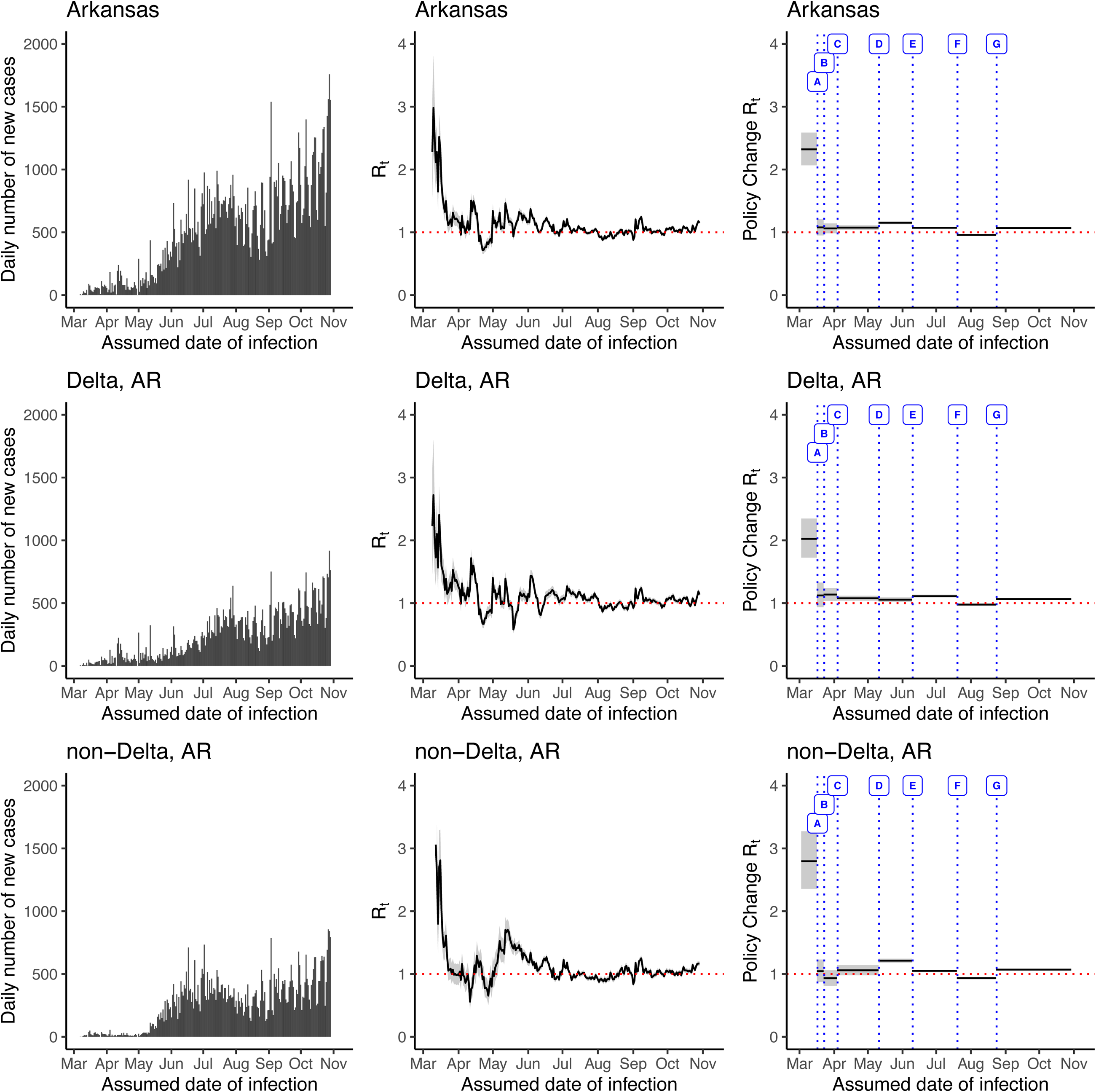
The daily number of incidence (left panel), time-varying reproduction number (*R_t_*) (middle panel), and *R_t_* per policy change (right panel) in Arkansas, USA, March 6 – November 7, 2020 (date of report), estimated using the instantaneous reproduction number method implemented in the ‘EpiEstim’ package. A = Schools closed; B = Restricted gatherings to 10 people or fewer; C = Required businesses, manufacturers, construction companies, and places of worship to implement social distancing protocols; D = Phase One reopening of restaurants, dine-in operations may continue; E = Governor announced that Phase 2 of reopening would begin on Jun 15, 2020, allowing restaurants and businesses to operate at two-thirds capacity; F = Required use of face coverings/masks in public; G = Schools reopened for in-person instruction

At the beginning, the PC*R_t_* estimates were high in Arkansas and both the Delta and non-Delta regions. The PC*R_t_* estimates declined statewide (median *R_t_* difference percentage: -53.56%, 95% CrI, -53.1%, -54.1%) and both Delta (-44.56%, 95% CI, -43.4%, -45.8%) and non-Delta regions (-62.67%, 95%, -62.4%, -63.0%) after schools closed on March 17^th^. The PC*R_t_* estimate remained stable statewide and in the Delta region when gatherings were restricted to 10 individuals or fewer on March 23^rd^, but declined by -10.81% (95% CrI, -26.9%, +8.35%) to below 1 in the non-Delta region. The PC*R_t_* estimates increased statewide (+6.68%; 95% CrI, +5.58%, +7.75%) and the non-Delta region (+14.29%; 95% CrI, -5.14%, +23.68%) after May 11^th^, when restaurant dine-in operations could resume. Both regions (Delta region: -12.08%; 95% CrI, -11.9%, -12.3%; Non-Delta region: -10.97%; 95% CrI, -10.6%, -11.3%), as well as Arkansas as a whole (-10.64%; 95% CrI, -10.60%, -10.70%), saw a decrease in the PC*R_t_* estimate when face masks were required in public beginning on July 20^th^. There was an increase in the PC*R_t_* estimates statewide (+11.56%; 95% CrI, +9.88%, +13.27%) and both regions (Delta region: +9.07%; 95% CrI, +6.85%, +11.18%; Non-Delta region: +14.51%; 95% CrI, +12.3%, +16.7%) after August 24^th^, when schools reopened with in-person instruction.

From March 6 to November 7, 2020, Kentucky’s daily incidence data showed a steady increase (**Figure 2**). In Kentucky, the 7-day sliding window *R_t_* estimate was high in March and decreased in April. The *R_t_* estimate had peaks that stayed around 1 and by the end of the study its median was 1.10 (95% CrI, 1.08, 1.12). Both regions (Appalachian and non-Appalachian) demonstrated an extensive community transmission of SARS-CoV-2, with a median 7-day sliding window *R_t_* larger than 1. The Appalachian and non-Appalachian regions’ median 7-day sliding window *R_t_* estimates were 1.07 (95% CrI, 1.04, 1.11) and 1.11 (95% CrI, 1.09, 1.14), respectively, at the end of the study.

**Figure 2.**
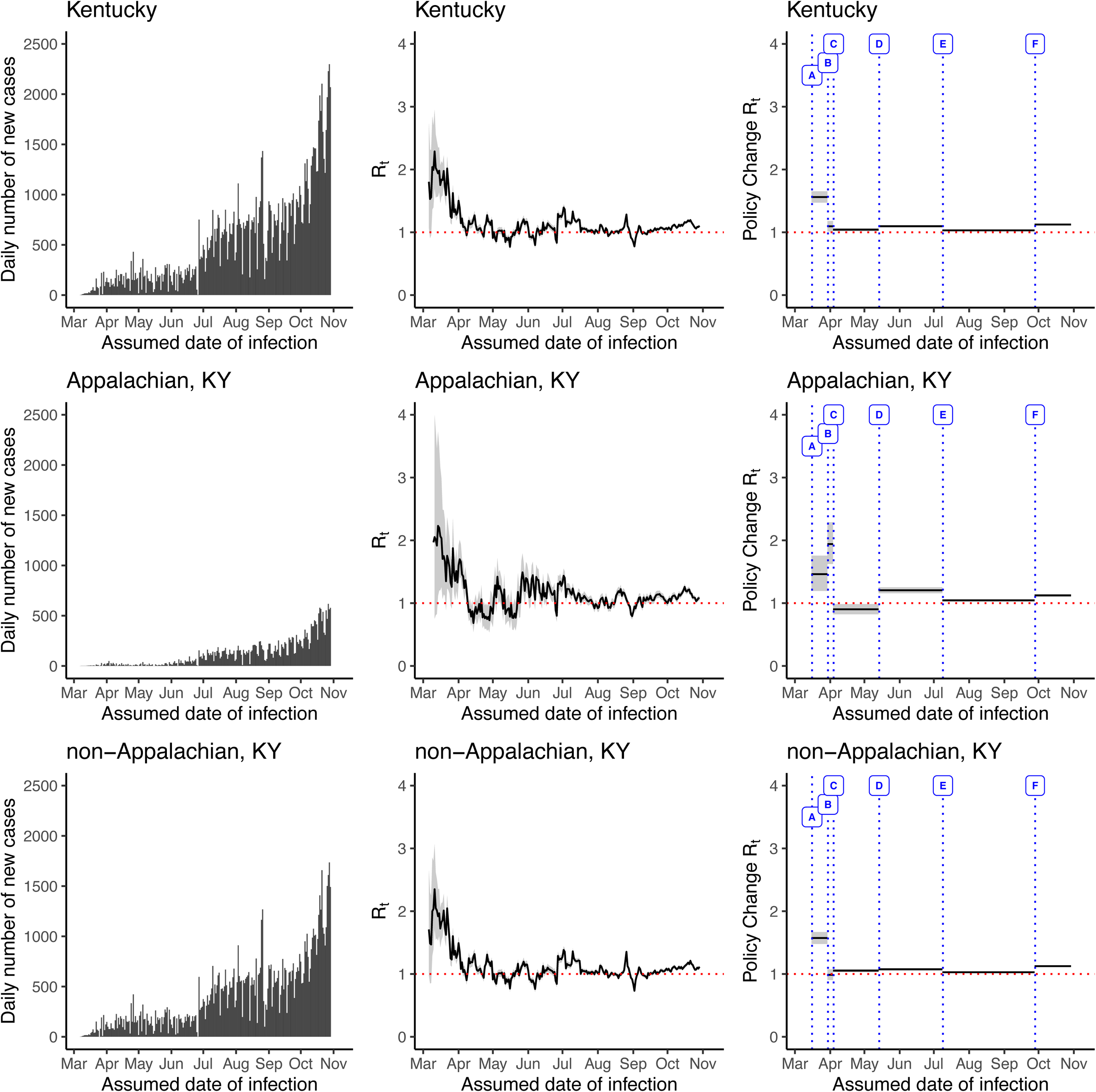
The daily number of incidence (left panel), time-varying reproduction number (*R_t_*) (middle panel), and *R_t_* per policy change (right panel) in Kentucky, USA, March 6 – November 7, 2020 (date of report), estimated using the instantaneous reproduction number method implemented in the ‘EpiEstim’ package. A = School closure and restaurants cease in-person dining; B = Order issued to restrict out-of-state travel; C = Adopted on a voluntary basis the new guidance from the U.S. Centers for Disease Control and Prevention (CDC) recommending that people wear cloth masks in some situations; D = Groups of 10 people or fewer may gather; E = Required use of face coverings/masks in public; F = Schools reopened with in-person instruction

The PC*R_t_* estimates were high among Kentucky and both regions, as the pandemic began spreading through the states. Out-of-state travel restrictions were issued on March 30^th^, decreasing the PC*R_t_* estimate statewide and in the non-Appalachian region, yet PC*R_t_* increased in the Appalachian region (+32.85%; 95% CrI, +30.3%, +35.8%). The PC*R_t_* estimate decreased to below 1 in the Appalachian region (-53.51%; 95% CrI, -45.16%, -61.2%) and remained stable in the entire state, after April 4^th^, when the state adopted on a voluntary basis guidance from the Centers for Disease Control and Prevention (CDC) recommending that individuals wear cloth masks in some situations. The PC*R_t_* estimates statewide (+5.19%; 95% CrI, +4.47%, +5.91%) and both regions (Appalachian region: +33.46%; 95% CrI, +20.7%, +46.8%; Non-Appalachian region: +1.93%; 95% CrI, +1.3%, +2.51%) increased after gatherings of 10 or less were allowed on May 14^th^. The PC*R_t_* estimates decreased to near 1 statewide (-5.93%; 95% CrI, -4.31%, - 7.65%) and both regions (Appalachian region: -13.34%; 95% CrI, -11.5%, -15.2%; Non-Appalachian region: -4.39%; 95% CrI, -2.56%, -6.33%) beginning on July 9^th^, with the executive order requiring face coverings in public. There was an increase in the PC*R_t_* estimates statewide (+8.97%; 95% CrI, +8.86%, +9.08%) and both regions after September 28^th^, when schools reopened with in-person instruction (Appalachian region: +7.49%; 95% CrI, +7.48%, +7.51%; Non-Appalachian region: +9.39%; 95% CrI, +9.23%, +9.56%).

## DISCUSSION

The purpose of this paper was to estimate and compare state and county-level *R_t_* trajectories of COVID-19 epidemics in Arkansas and Kentucky, focusing on differences between urban and rural areas. The implementation of preventative and relaxation measures impacted case burden and the direction of the *R_t_* trajectories. We observed decreased *R_t_* estimates when facial coverings were mandated, changing by -10.64% in Arkansas and -5.93% in Kentucky from the previous policy interval.

This paper uses *R_t_* to examine the COVID-19 transmission over several months, as well as examine how it varied by public health interventions and policy changes. The *R_t_* estimates provided public health policy makers near-real time indicators of the trajectory of the epidemic and whether their public health interventions were able to put the epidemic under control.

Several studies have examined the *R_t_* estimates with respect to policy and interventions and used *R_t_* estimates as predictive models and quantitative measures of epidemic growth or decline.^31–33^ Here, the *R_t_* trajectories of Arkansas and Kentucky differed among rural and urban areas, increasing or decreasing, depending on the implementation of preventative and relaxation measures. The *R_t_* will be useful as the pandemic progresses to inform policymakers and public health professions of the direction of potential outbreaks, assisting in preventing health care surges and implementing more preventative measures and policies. For example, both Kentucky and Arkansas implemented mandated facial coverings or masks in July, 2020, which was reflected by a decrease in COVID-19 transmission.

Our study sought to further examine if differences in COVID-19 transmission occurred among location, specifically urban versus rural, since we observed that the role of population size in counties has had a less significant effect on the spread of COVID-19. One study examined trends in the distribution of COVID-19 hotspot counties and found that more hotspot counties were occurring in the southern states of the US during summer months in 2020.^34^ This followed the trend and wave progression that occurred in the US, hitting the large metropolitan areas first, followed by spread in the Southern region and then in the Mid-West region. Another study found that that many of the less vulnerable counties that had a low Social Vulnerability Index had slightly higher average incidence and death rates early in the pandemic, and as the pandemic progressed, the trends crossed, with many of the most vulnerable counties facing higher rates.^35^ Many of the urban metropolitan areas and cities were impacted first, before spreading to the rural areas. This may be due to the linkage of metropolitan areas, through social, economic, and commuting relationships.

Arkansas, one of eight states in the US that did not implement a stay at home or lockdown order, lacked the immediate response, as seen by other states, could explain the higher *R_t_* estimate, as it was at 2 or higher at the beginning of the pandemic.^36^ Arkansas had 22 cases before the first preventative measure, the closing of schools on March 17^th^, was implemented.

Additionally, the only time the PC*R_t_* estimate was below 1 was when face coverings were implemented in July, demonstrating a decrease in COVID-19 transmission. One of the biggest drivers in COVID-19 transmission in Arkansas was the poultry plant outbreaks that occurred among employees and spread through community transmission.^37^ In Lincoln County, Arkansas, many COVID-19 cases were attributable to the correctional facility outbreak, rather than community transmission.^38^ Additionally, there was an increase in mass testing at the correctional facility in Lincoln County, which could explain the large peaks in *R_t_* estimates that we observed.^39^ One study conducted among a correctional facility in Arkansas observed that if testing for COVID-19 was only among symptomatic individuals, then fewer cases would have been detected, allowing for a greater transmission of disease to occur.^40^

At the beginning of the pandemic, many states in the South and Midwest of the US observed increased COVID-19 infection rates, yet Kentucky’s rate was notably low.^41^ Kentucky took a very conservative method in their approach, as was observed by the policies and measures implemented, to slow the transmission of COVID-19. A decrease in COVID-19 transmission was seen in the Appalachian region, when the state adopted the guidance from the CDC recommending that people wear cloth masks in some situations and when Kentucky passed an executive order requiring face coverings in public. The Kentucky Appalachian region has high rates of comorbidities, especially respiratory diseases due to the coal industry, but saw an increase in mask wearing when required.^42^ In Jefferson, Shelby, and Warren Counties in Kentucky, a decrease in PC*R_t_* was observed in transmission towards the beginning of the pandemic, when an order was issued to restrict out-of-state travel. This decrease in transmission may have been due to less travel that occurred across state lines, as Warren County is near the Tennessee border and Nashville, the Tennessee capital, and Jefferson and Shelby Counties border Indiana, and is near Cincinnati in Ohio.^43^

During the study period, both Arkansas and Kentucky maintained a median *R_t_* above 1. These two states had different political parties in charge of the governor’s office in 2020, yet share similar cultural and heritage histories. Additionally, within both of these states, we found that mandated face coverings were associated with a decreased *R_t_* estimate and reopening of schools were associated with an increased *R_t_* estimate. There are a few similarities of *R_t_* estimates and PC*R_t_* estimates among these two states on a statewide level, which may suggest underlying factors, such as COVID-19 variants and pathology, rather than social determinants of health.

However, once we examine regional level, and even county level, we find both similarities (decreased COVID-19 transmission with mandated face coverings) and differences (increased COVID-19 transmission with gatherings of 10 or less allowed). The findings of this study among two similar southern states also relates to many other regions. Among different regions in the US, face coverings mandates and reopening of schools also showed a decreased and increased, respectively, of COVID-19 transmission. In the Western states (North Dakota, Montana, and Wyoming), it was found that the *R_t_* estimate decreased following a face covering mandate.^44^ An increase in COVID-19 transmission was observed in South Carolina following the reopening of schools (15.3%).^45^

While the *R_t_* differed among rural and urban areas at the beginning of the pandemic, as the pandemic progressed, the *R_t_* was similar across the urban and rural counties in both states. Although population size has been found to have a less significant effect on COVID-19 spread than hypothesized at the early pandemic, it is still important to discuss the disparities that occur between rural and urban locations and the implications the pandemic has on rural locations. Rural areas have had lower testing rates, as well as poorer health care infrastructures to handle cases.^46^ Rural health care and public health systems are more vulnerable and have struggled to respond to the COVID-19 crisis.^47^ Additionally, most healthcare systems do not have the capacity to handle surges in cases, and only one percent of the nation’s intensive care unit beds are located in rural areas.^48^ Many care and patient populations are different in rural communities and it is an important aspect to understanding the spread of COVID-19. Although policy and preventative measures are statewide, it does show differences among rural and urban communities. One study found that rural Americans were less likely than urban Americans to follow most recommended COVID-19 prevention behaviors.^49^

There were several limitations in this study. One limitation was the lack of data on superspreading events that occurred in each state (for example, within prisons^50^ and nursing homes,^51^ as well as in religious settings, schools and sport camps, and social events^52^). The lack of testing data, as well as hospitalization data, by county level may lead to testing bias. This would have provided further insight into the rural and urban disparities that may be present.

Many of the counties located in both Arkansas and Kentucky contained large prison populations. The counties of Lincoln, Arkansas and Elliot, Kentucky, both contain county correctional facilities and prisons.^38, 53^ The reason for the unstable *R_t_* in these counties may stem from disease amplification in prison outbreaks rather than community spread. However, it is difficult to pinpoint certain related outbreaks, and there is limited county-level data specific to correctional facilities. Additionally, there were 1,755 unknown county-level cumulative cases in Arkansas. These cases were included in our state-level data analysis, but they were excluded from the Delta, non-Delta, and county-level hot spots analyses. Kentucky had all county-level data and all reported cases were used in all analyses.

This study observed that both Arkansas and Kentucky, as well as the respective regions, had an extensive spread of COVID-19, since both states maintained a median *R_t_* above 1. The direction of the trend of the *R_t_* estimates were reflected by the implementation of preventative measures and their subsequent relaxation as the pandemic progressed. This study was able to examine the changing transmission potential of COVID-19 over time in rural and urban areas in two socio-demographically similar Southern states. Further research is needed to examine the rural and urban differences in the spread of the COVID-19 pandemic in the US.

## Data Availability

The dataset used in this manuscript is publicly available at the New York Times GitHub data repository at https://github.com/nytimes/covid-19-data.

## Acknowledgments

The preliminary version of this project has its origin from an MPH class group project (EPID 7135 Epidemiology of Infectious Diseases). The authors would like to thank Ar’reil Smithson, MPH, Diane Martinez-Piedrahita, MPH, Holly Richmond-Woods, DVM, MPH, and Terrance D. Jacobs, MPH for their participation in the class group project. The authors would like to thank Aubrey Dayton-Kehoe, MPH, for sharing her R code with us.

## Appendix A. Management of Negative Incident Case Counts

If there were any negative daily case counts in the data (i.e., when public health agencies made corrections to their cumulative case counts at a specific date or dates), they were identified and adjusted by changing the negative cases counts to zero and correcting the daily case counts on previous days such that the cumulative case counts of the previous days would not exceed the cumulative case counts of the day that reported negative daily case counts. We adjusted the negative daily case counts at both state and county levels. These data management steps were done in R 4.0.3.

## Appendix B. The Instantaneous Reproduction Number Rt (“EpiEstim” package)

Time-varying reproduction number, denoted as *R_t_*, was estimated using the R package EpiEstim version 2.2-3; the method used was the instantaneous reproduction number method with parametric definition of the serial interval.^1^ This measure was defined by Cori et al.^1^ as the ratio between *I_t_*, the daily number of new case at the time *t*, to the total infectiousness of all infected individuals at the time *t*. Λ_t_ is mathematically interpreted as, 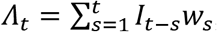, that is, the sum of the total infected individuals up to *t-1*, weighted by the infectivity function *w_s_*, a probability distribution that describes the average infection and is typically expressed by the serial interval distribution. Hence, the daily new case counts at time *t* is Poisson-distributed with a mean of 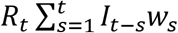. Contingent on the number of daily new cases in previous time points, *I_0_*, …, *I_t-1_*, and given the reproduction number *R_t_*, the likelihood of the daily number of new cases *I*_t_ is as follows: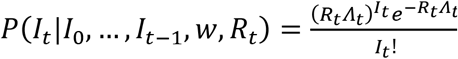 where the total infectiousness of infected individuals at time 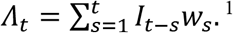. In other words, 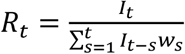 can be interpreted as the average number of secondary cases infected at time *t* by an infectious person if conditions remain the same. Since this formulation can result in a highly variable *R_t_* estimate over time, the method of instantaneous reproduction number as implemented in EpiEstim assumes that the *R_t_* is constant over a given time frame of size *τ* ending at time *t*. This will enable a precise estimate to be estimated with limited variability. Given that transmissibility is assumed to be constant over a period of time, from *(t-τ+1)* to *t*, and is denoted by a reproduction number, *R_t,τ_*, the possibility of the number of daily new cases during the time period, from *I_(t-τ+1)_* to *I_t_*, is contingent on daily number of new cases prior to the time period, i.e., from *I_0_*, to *I_(t-τ)_.*^1^ Cori et al.^1^ derived an analytical expression of the posterior distribution of Rt and thus estimated its median, the variance, and the 95% credible interval by using a Bayesian framework with a Gamma-distributed prior to *R_t,τ_*,

In this paper, we estimated *R_t_* using both a 7-day sliding window and non-overlapping time periods specified by the dates of policy changes. The data was analyzed using EpiEstim version 2.2-3.^1^

## Appendix C. Regional County-level Hot-Spot Analyses

We chose four Arkansas counties for further analysis: Washington County, Benton County, Lincoln County, and Yell County. We combined Washington County (where Fayetteville is) and Benton County (second most populous county in Arkansas) together for analysis because these two counties are geographically next to each other and large metropolitan areas, and therefore the frequency of daily communication is high.

Washington County and Benton County’s daily number of new cases revealed two major surges in June and September (**Supplementary Figure 1**). The 7-day sliding window *R_t_* estimates in Washington County and Benton County was low at the beginning of the pandemic, either near 1 or below 1, reaching a peak of an *R_t_* estimate of 2 in mid-May, before steadily staying around 1 and increasing to above 1 in a peak occurring in mid-August. At the end of the study, the median *R_t_* estimate was 1.30 (95% CrI, 1.23, 1.36). For Lincoln County, a major outbreak occurred in April. Lincoln County started with peaks with the 7-day sliding window *R_t_* estimates nearing 3, and then decreasing to below 1 and then having peaks and troughs as the pandemic progressed. At the end of the study, the median *R_t_* estimates for Lincoln County was 0.66 (95% CrI, 0.42, 0.99). For Yell County, the major surge of new cases occurred between June and July. Yell County had peaks in the beginning with the 7-day sliding window *R_t_* estimates above 3, before having smaller peaks and troughs that remained around 1. At the end of the study, the median *R_t_* was 1.24 (95% CrI, 0.90, 1.65). Except Lincoln County, the rest of the three counties demonstrated an extensive community transmission of SARS-CoV-2 transmission with the median 7-day sliding window *R_t_* more than 1.

Among the policy change *R_t_* plots, Washington and Benton counties followed a similar pattern that was seen for the overall state of Arkansas. There was a decrease in the policy change *R_t_* estimate when gatherings were restricted to 10 or fewer on March 23^rd^ (median *R_t_* difference percentage: -30.66%; 95% CrI, -28.0%, -34.0%), but increased (+68.04%; 95% CrI, +15.7%, +147.6%) again to above 1 when businesses, manufacturers, construction companies and places of worship were required to implement social distancing protocols on April 4^th^. There was another decrease (-9.77%; 95% CrI, -8.24%, -11.25%) in the policy change *R_t_* estimate to below 1 when face coverings and masks were required in public on July 20^th^, but it increased (+22.08%; 95% CrI, +19.5%, +24.7%) once schools reopened for in-person instruction on August 24^th^. Lincoln County did not have a clear pattern in terms of policy change *R_t_* estimates. For example, the largest decline in the policy change *R_t_* estimate was when the state allowed for the reopening of restaurants, with the continuation of dine-in operations on May 11^th^ (-46.87%; 95% CrI, -25.5%, -62.4%). There was also a slight, but not significant increase in the policy change *R_t_* estimate (+6.15%; 95% CrI, -10.4%, +23.9%) when face coverings and masks were required, and also a decline below 1 in the policy change *R_t_* estimate (-22.91%; 95% CrI, -15.7%, -29.9%) when schools reopened again with in-person instruction. However, Lincoln County contains a large correctional facility that may be driving the transmission in a congregate setting instead of community transmission.^2^ Yell County did not have a significant decrease when face coverings and masks were required (-2.31%; 95% CrI, -14.7%, +13.6%), nor a significant increase when schools reopened with in-person instruction (+4.18%; 95% CrI, -12.4%, +23.8%). We chose four Kentucky counties for further analysis: Jefferson County, Shelby County, Elliot County, and Warren County. Jefferson County is where Louisville is and Shelby County is its suburb. We combined Jefferson County and Shelby County together for analysis because these two counties are geographically next to each other therefore human mobility between the two counties is high. The Jefferson County and Shelby County’s daily number of new cases reveal steady increase (**Supplementary Figure 2**), and the median 7-day sliding window *R_t_* estimates was 1.05 (95% CrI, 1.01, 1.09) for the assumed date of infection. For Elliott County, the major outbreak happened in October. At the end of the study, the median 7-day sliding window *R_t_* estimate Elliott County was 0.94 (95% CrI, 0.70, 1.24). For Warren County, it had a steady increase with the major surges of new cases in May. At the end of the study, the median 7-day sliding window *R_t_* was 1.23 (95% CrI, 1.11, 1.35). All four counties demonstrated an extensive community transmission of SARS-CoV-2 transmission with the median 7-day sliding window *R_t_* very close to and more than 1.

Jefferson County and Shelby County in Kentucky followed a similar pattern that was seen for the overall state of Kentucky for the policy change *R_t_* plots. There was a decrease in the policy change *R_t_* estimate when out of state travel was restricted on March 30^th^ (median *R_t_* difference percentage: -57.09%; 95% CrI, -48.1%, -64.5%), but increased again to above 1 on April 4^th^ (+59.92%; 95% CrI, +32.0%, +95.6%), when the state adopted on a voluntary basis the new guidance from the CDC recommending that individuals wear cloth masks in some situations. The policy change *R_t_* estimate decreased when groups of 10 could gather (-30.66%; 95% CrI, -28.0%, -34.0%), remained the same when face coverings and masks were required in public (+1.79%; 95% CrI, -0.08%, +3.64%), and increased once schools reopened for in-person instruction (+6.21%; 95% CrI, +5.8%, +6.63%). Sustained low transmission was found in Elliot County even after facemasks was mandated in July. However, the real change was the reopening of schools with in-person instructions. An outbreak in October ensued with a policy change *R_t_*>1. Warren County had a policy change *R_t_* estimate of 1.52 (95% CrI, 0.59, 3.12) at the beginning of the pandemic, and 1.97 (95% CrI, 1.31, 2.81) after schools were ordered to close (+29.33%; 95% CrI, -42.9%, +253.7%). The policy change *R_t_* estimate decreased to below 1, when the order was issued to restrict out-of-state travel (-54.23%; 95% CrI, -14.90%, -76.2%). However, there was no significant increase when facial coverings were mandated (+2.37%; 95% CrI, -5.07%, +10.0%) and increased to above 1 when schools reopened with in-person instruction (+11.59%; 95% CrI, +7.45%, +16.83%).

We only present the 1-week sliding window *R_t_* and policy change *R_t_* with y-axis ranging between zero to four because anything larger than 4 with small data size is not reliable. Given that the *R_0_* of COVID-19 is accepted to be around 3 with some viral variants with a slightly higher *R_0_*^3, 4^ and that *R_t_* would be < *R_0_*, it is reasonable to assume 0 < *R_t_* < 4. The same method applied to the starting date of *R_t_* estimation: the R package EpiEstim has a default *R*_t_ estimation (mean value of the prior) of 5 and the 95% CrI is wide with limited daily case count. Therefore, in the county-level hot spot analyses, we only recorded the *R_t_* estimation from the month that those locations experienced first major local outbreaks (e.g., for Elliot County since it had a small daily case count in the beginning of the pandemic until August, 2020).

## Appendix D. Statewide Hospitalization Analyses

As a sensitivity analysis, we examined the 3-day moving averages of statewide hospitalization data from Arkansas and Kentucky, shifting 12 days for the hospitalization data to get the assumed infection date (**Supplementary Figure 5**).

Arkansas had very low daily number of COVID-19 hospitalizations who were infected in March-April 2020, with a few dates reporting zero or no cases. The number continued to steadily rise, before reaching its largest peak in July (assumed date of infection). Because of multiple zero entries in the Arkansas hospitalization data in March-April 2020, we only presented *R_t_* estimate starting on April 29 (assumed date of infection). The *R_t_* estimate in Arkansas started below 1 in late April, and reached its highest estimate in mid-May. The *R_t_* estimate mainly was above 1, with a few points falling below 1, before steadily staying around 1.

Kentucky had a very large daily number of COVID-19 hospitalizations who were infected in March-April 2020. The daily number of COVID-19 hospitalizations declined until July, when two peaks of hospitalized cases were assumed to be infected, and declining again until rising at the end of September and the end of October (assumed date of infection). The *R_t_* estimate among Kentucky hospitalizations mostly remained below 1, before reaching an *R_t_* estimate above 2 in July (first peak), followed by another peak with an *R_t_* estimate of about 1.5, and ending with an *R_t_* estimate under 1 in August. There were two more peaks that had an *R_t_* estimate above 1, at the end of September and end of October.

## Appendix E. Converting the power-law relationship between cumulative case count and population size to a linear relationship between log-transformed per capita cumulative case count and log-transformed population size

As in Fung et al.,^5^ we explored the power-law relationship between the cumulative case count of COVID-19 (C) and their Census-estimated population size (N), C∼N^g^ (where g is the exponent), as follows:

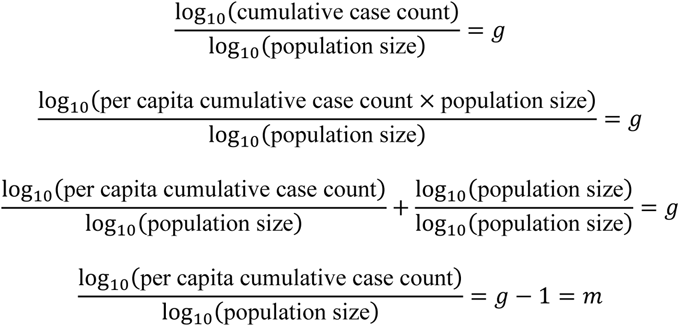

Where *m* is the slope of the regression line between the log-transformed per capita cumulative case count and the log-transformed population size.

Per capita cumulative incidence would be exactly proportional to population size, and there was no heterogeneity of per capita cumulative incidence across geographic units of different population sizes if m=0 (i.e., g=1). Geographical units with lower population sizes would have a higher per capita cumulative incidence if m<0 (i.e., g<1) and lower per capita cumulative incidence if m>0 (i.e., g>1).

**Supplementary Figure 6** presents a linear regression relationship between the log10-transformed per capita cumulative case number and the log10-transformed population size for a total of 195 counties in Arkansas and Kentucky. Each panel in the figure corresponds to an assessed date, May 7, July 7, September 7, and November 7, 2020, respectively. The non-Appalachian region had a positive slope consistently at four different assessed dates (0.29 (95% CI, 0.07, 0.52), 0.24 (95% CI, 0.07, 0.41), 0.13 (95% CI, 0.02, 0.23), 0.10 (95% CI, 0.02, 0.18), [**Supplementary Table 7**]), indicating that counties with a higher population size would have a higher per capita cumulative case number. However, for other regions, there was no evidence to reject the hypothesis of homogeneity of per capita cumulative incidence across counties of different population sizes, except the Appalachian region had a positive slope, 0.39 (95%CI, 0.10, 0.68), on July 7, 2020.

At the beginning of the pandemic, it was hypothesized that population density and size was the main factor for the rapid spread of COVID-19, especially in cities such as Wuhan, New York, and Milan.^6^ However, many high-population, high-density cities, including Hong Kong^7^ and Seoul,^8^ were able to successfully limit both COVID-19 cases and deaths with population-level interventions, such as mask wearing, social distancing, and contact tracing. One study found that denser places were not linked to higher infection rates and were associated with lower COVID-19 death rates.^9^ Another study found that larger metropolitan size was linked to higher infection and mortality rates over time, however, during the same time period, higher population density was linked to lower infection and mortality rates.^10^ Although our study examined the differences in *R_t_* among urban and rural locations to determine if rurality had a significant role in the spread of COVID-19, we did not examine population density; however, we did find that population size was not a factor, with the exception of the non-Appalachian region. This supports the hypothesis that population size has not played a major role in the spread of COVID-19.

Another study found that population outflow and migration in Wuhan contributed to the spatial-geographical spread of COVID-19 in that region.^11^ We observed higher levels of COVID-19 transmission in the hot spot analyses of the metropolitan counties of Washington and Benton in Arkansas, as well as the metropolitan counties of Jefferson and Shelby in Kentucky. This might be due to a larger number of commuters from surrounding counties that travel for work or school; however, further research is needed to investigate if the COVID-19 transmission in these metropolitan counties was due to commuters or the population outflow and inflow since this study did not examine the spatial-geographical spread among communities and counties.

**Supplementary Figure 1.**
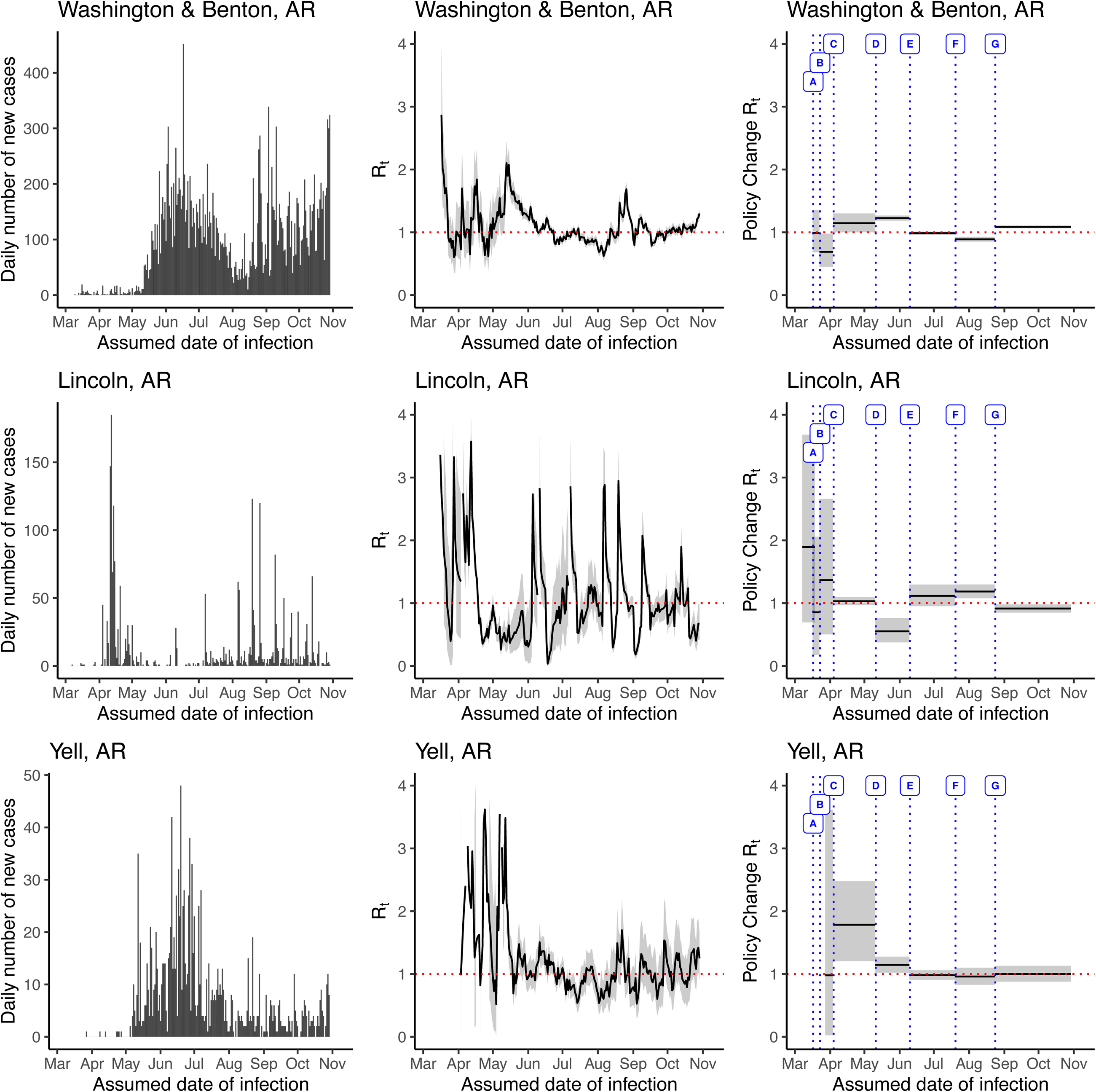
The daily number of incidence (left panel), time-varying reproduction number (*R_t_*) (middle panel), and *R_t_* per policy change (right panel) in Washington and Benton, Lincoln and Yell, Arkansas, USA, March 6 – November 7, 2020 (date of report), estimated using the instantaneous reproduction number method implemented in the ‘EpiEstim’ package. Legend: A = Schools closed; B = Restricted gatherings to 10 people or fewer; C = Required businesses, manufacturers, construction companies, and places of worship to implement social distancing protocols; D = Phase One reopening of restaurants, dine-in operations may continue; E = Governor announced that Phase 2 of reopening will begin on Jun 15, 2020, allowing restaurants and businesses to operate at two-thirds capacity; F = Required use of face coverings/masks in public; G = Schools reopened for in-person instruction

**Supplementary Figure 2.**
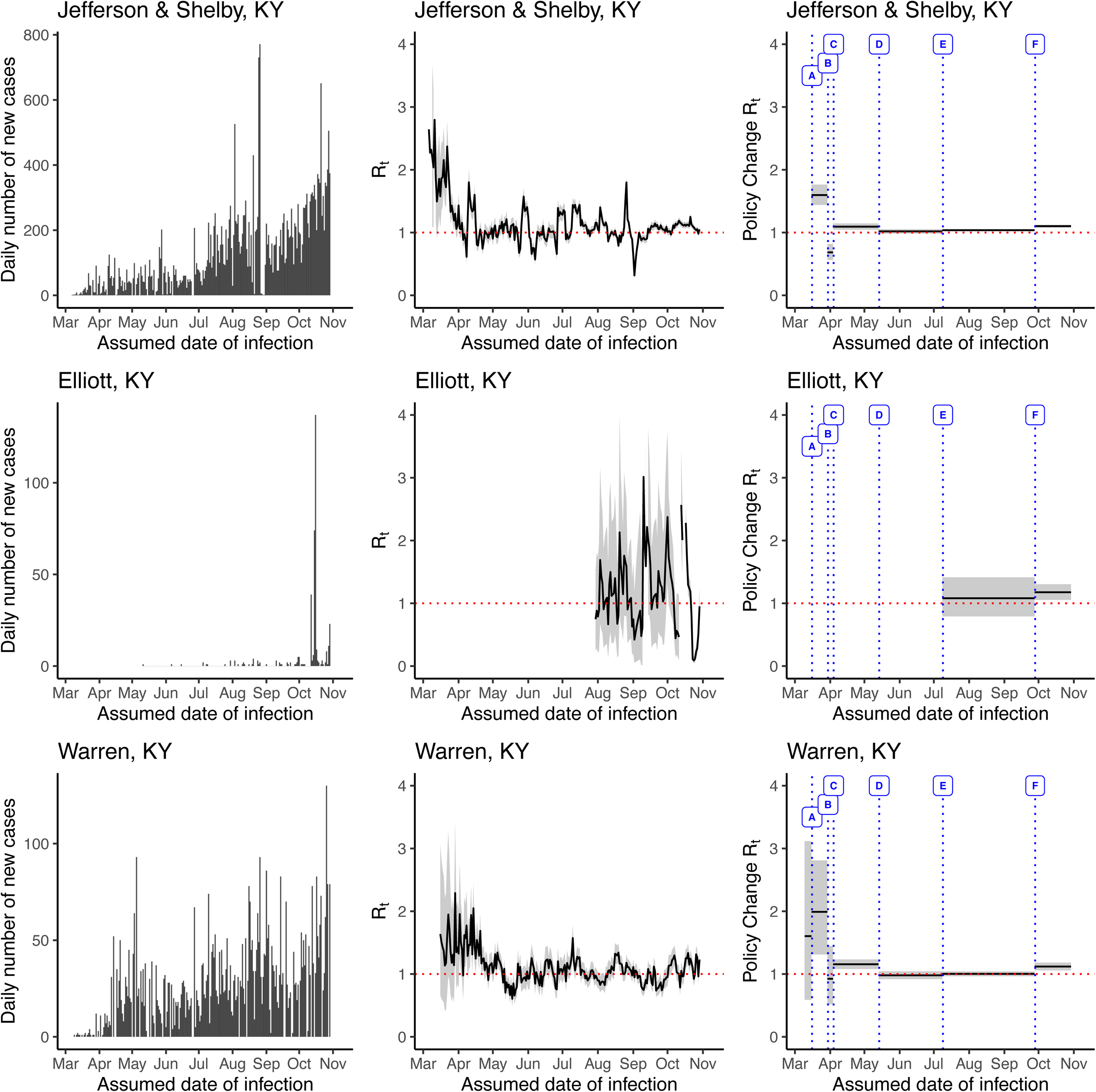
The daily number of incidence (left panel), time-varying reproduction number (*R_t_*) (middle panel), and *R_t_* per policy change (right panel) in Jefferson and Shelby, Elliott, and Warren, Kentucky, USA, March 6 – November 7, 2020 (date of report), estimated using the instantaneous reproduction number method implemented in the ‘EpiEstim’ package. Legend: A = School closure and restaurants cease in-person dining; B = Order issued to restrict out-of-state travel; C = Adopted on a voluntary basis the new guidance from the U.S. Centers for Disease Control and Prevention (CDC) recommending that people wear cloth masks in some situations; D = Groups of 10 people or more may gather; E = Required use of face coverings/masks in public; F = Schools reopened with in-person instruction

**Supplementary Figure 3.**
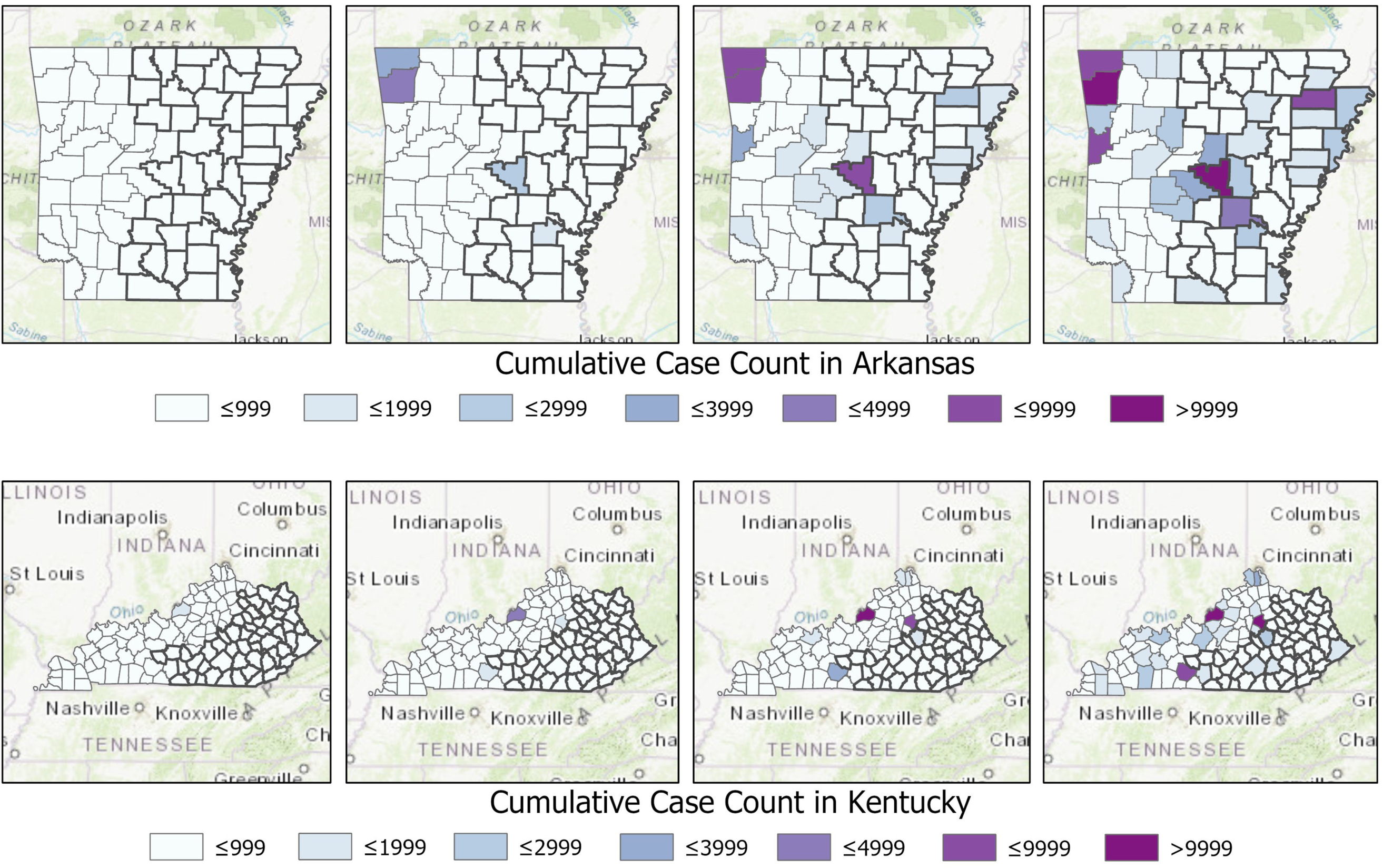
Maps of cumulative case count in Arkansas (top) and Kentucky (bottom) on May 7, July 7, September 7, and November 7, 2020 (date of report).

**Supplementary Figure 4.**
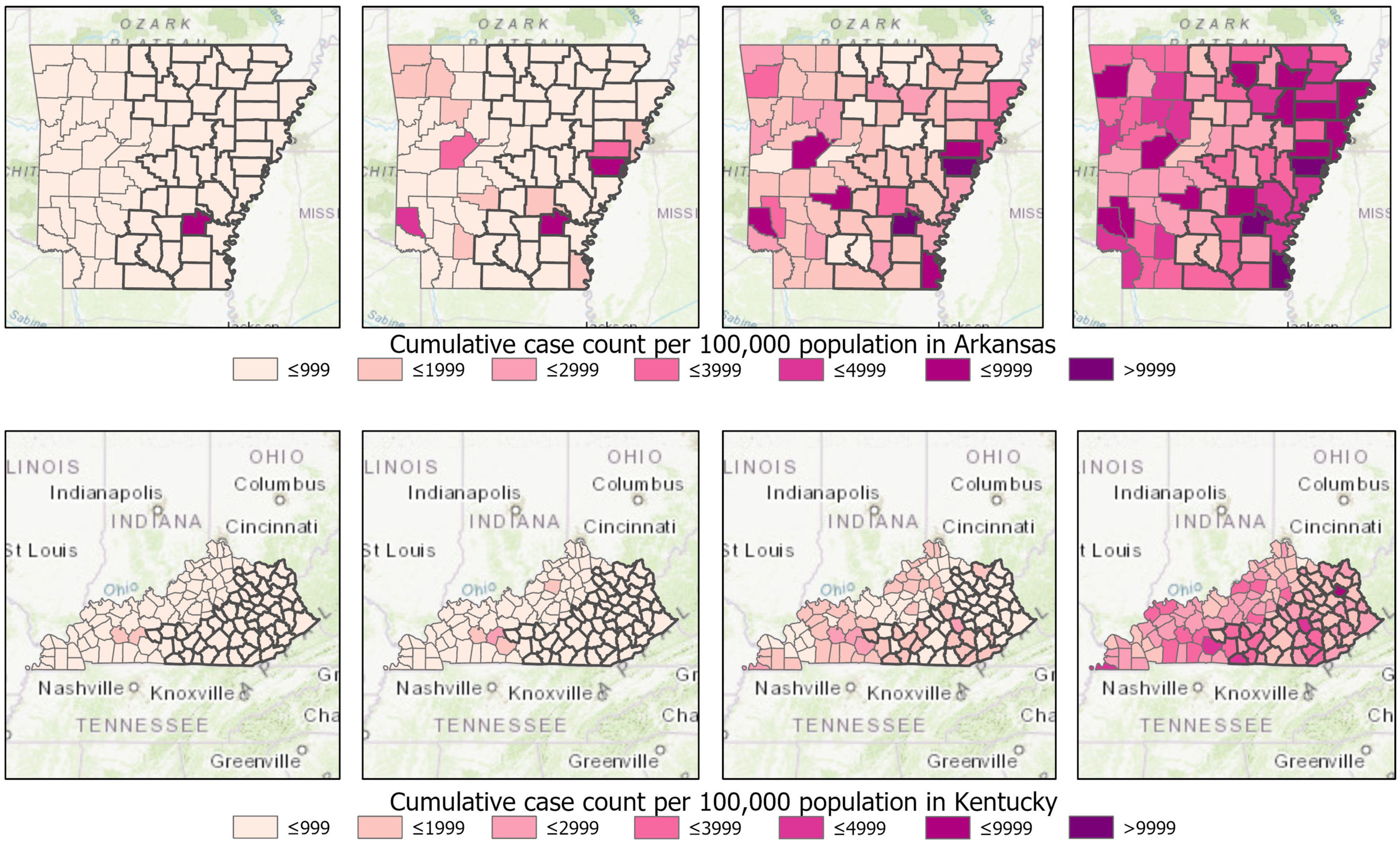
Maps of cumulative case counts per 100,000 population in Arkansas (top) and Kentucky (bottom) on May 7, July 7, September 7, and November 7, 2020 (date of report).

**Supplementary Figure 5.**
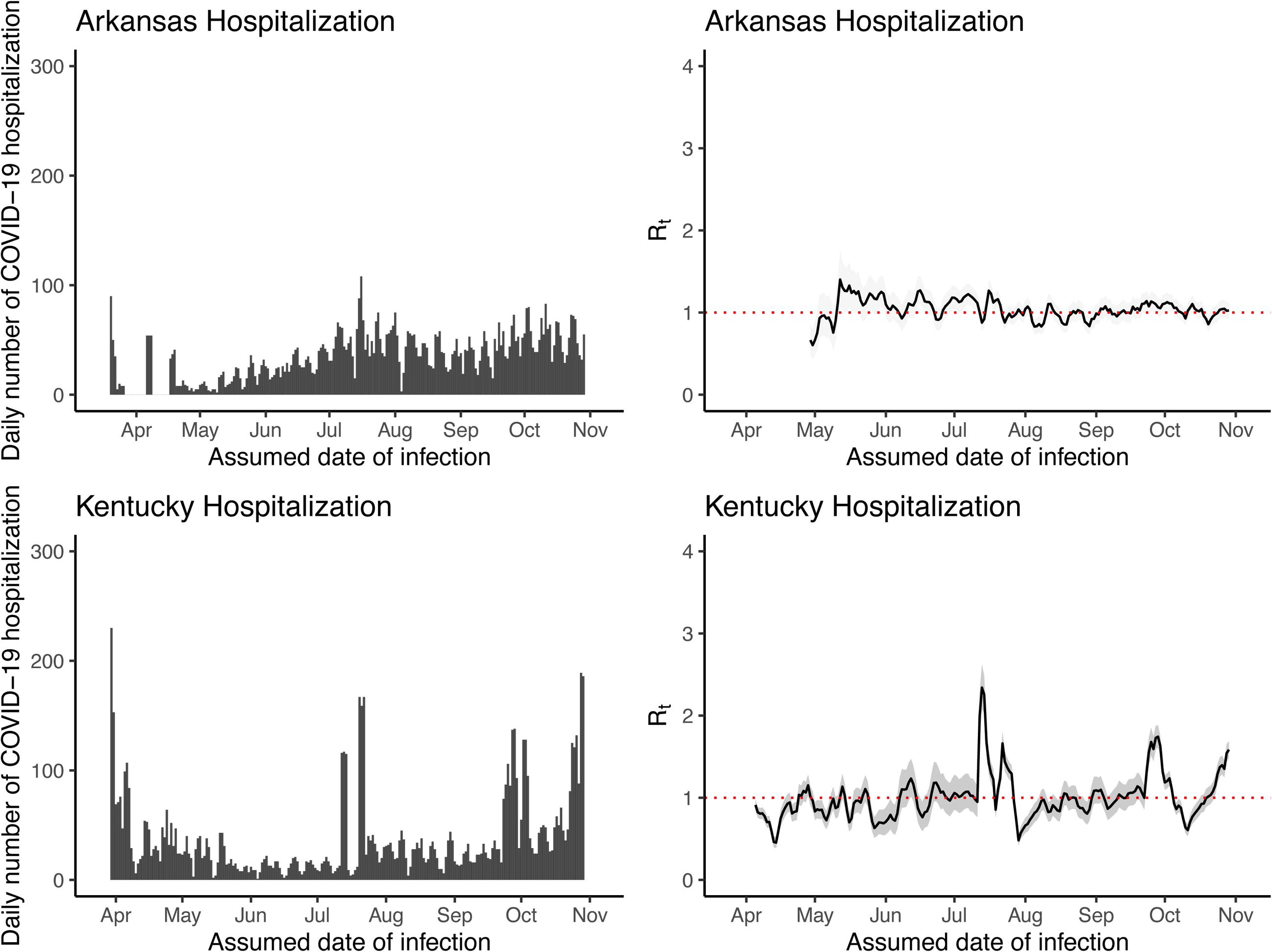
The daily number of COVID-19 hospitalizations (left panel), and *R_t_* estimates derived from hospitalization data (right panel) in Arkansas and Kentucky.

**Supplementary Figure 6.**
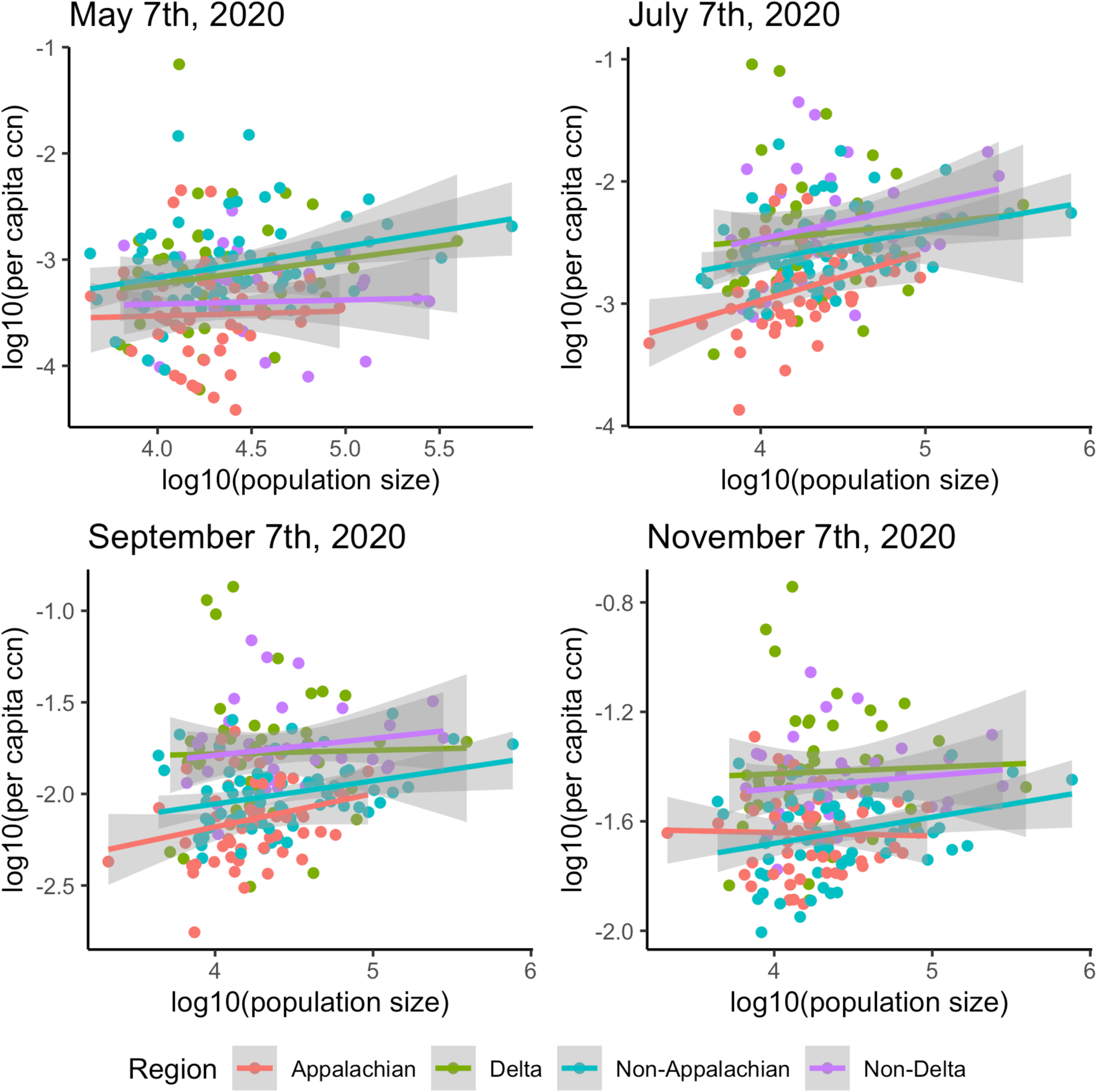
Linear regression between log10-transformed per capita cumulative case number (ccn) and log10-transformed population size by county, grouped in regions: Appalachian (red), Delta (green), Non-Appalachian (blue) and Non-Delta (purple), in the states of Arkansas and Kentucky on May 7^th^, July 7^th^, September 7^th^, and November 7^th^, 2020

**Supplementary Table 1.** Arkansas Counties by Region.

| **Region** | **Counties** |
| --- | --- |
| Delta Region (42 counties) | Arkansas, Ashley, Baxter, Bradley, Calhoun, Clay, Cleveland, Chicot, Craighead, Crittenden, Cross, Dallas, Desha, Drew, Fulton, Grant, Greene, Independence, Izard, Jackson, Jefferson, Lawrence, Lee, Lincoln, Lonoke, Marion, Mississippi, Monroe, Ouachita, Phillips, Poinsett, Prairie, Pulaski, Randolph, St. Francis, Searcy, Sharp, Stone, Union, Van Buren, White, Woodruff |
| Non-Delta Region (33 counties) | Benton, Boone, Carroll, Clark, Cleburne, Columbia, Conway, Crawford, Faulkner, Franklin, Garland, Hempstead, Hot Spring, Howard, Johnson, Lafayette, Little River, Logan, Madison, Miller, Montgomery, Nevada, Newton, Perry, Pike, Polk, Pope, Saline, Scott, Sebastian, Sevier, Washington, Yell |

**Supplementary Table 2.** Kentucky Counties by Region.

| **Region** | **Counties** |
| --- | --- |
| Appalachian Region (54 counties) | Adair, Bath, Bell, Boyd, Breathitt, Carter, Casey, Clark, Clay, Clinton, Cumberland, Edmonson, Elliott, Estill, Fleming, Floyd, Garrard, Green, Greenup, Harlan, Hart, Jackson, Johnson, Knott, Knox, Laurel, Lawrence, Lee, Leslie, Letcher, Lewis, Lincoln, McCreary, Madison, Magoffin, Martin, Menifee, Metcalfe, Monroe, Montgomery, Morgan, Nicholas, Owsley, Perry, Pike, Powell, Pulaski, Robertson, Rockcastle, Rowan, Russell, Wayne, Whitley, Wolfe |
| Non-Appalachian Region (66 counties) | Allen, Anderson, Ballard, Barren, Boone, Bourbon, Boyle, Bracken, Breckinridge, Bullitt, Butler, Caldwell, Calloway, Campbell, Carlisle, Carroll, Christian, Crittenden, Daviess, Fayette, Franklin, Fulton, Gallatin, Grant, Graves, Grayson, Hancock, Hardin, Harrison, Henderson, Henry, Hickman, Hopkins, Jefferson, Jessamine, Kenton, Larue, Livingston, Logan, Lyon, Marion, Marshall, Mason, McCracken, McLean, Meade, Mercer, Muhlenberg, Nelson, Ohio, Oldham, Owen, Pendleton, Scott, Shelby, Simpson, Spencer, Taylor, Todd, Trigg, Trimble, Union, Warren, Washington, Webster, Woodford |

**Supplementary Table 3.** Time-varying reproduction number in Arkansas estimated using non-overlapping time windows (median and 95% credible interval) and its change between each time window (median and 95% credible interval)

| Time window | Median Rt & 95% CrI | Median Rt difference percentage changes comparing with previous policy interval & 95% CrI |
| --- | --- | --- |
| Arkansas |  |  |
| Before policy A | 2.3174 (2.07, 2.59) |  |
| From A to B | 1.0761 (0.949, 1.214) | -53.56% ( -53.1%, -54.1%) |
| From B to C | 1.0610 (0.984, 1.142) | -1.49% (-1.72%, +2.12%) |
| From C to D | 1.0743 (1.04, 1.11) | +1.11% (-9.32%, +6.94%) |
| From D to E | 1.1515 (1.13, 1.18) | +6.68% (+5.58%, +7.75%) |
| From E to F | 1.0716 (1.06, 1.08) | -7.09% (-5.07%, -8.95%) |
| From F to G | 0.9571 (0.954, 0.97) | -10.64% (-10.60%, -10.70%) |
| Beyond G | 1.0680 (1.06, 1.08) | +11.56% (+9.88%, +13.27%) |
| Delta region, Arkansas |  |  |
| Before policy A | 2.0203 (1.73, 2.35) |  |
| From A to B | 1.1195 (0.934, 1.328) | -44.56% (-43.4%, -45.8%) |
| From B to C | 1.1355 (1.04, 1.24) | +1.31% (-15.6%, +20.0%) |
| From C to D | 1.0779 (1.04, 1.12) | -5.27% (-14.2%, +4.24%) |
| From D to E | 1.0543 (1.02, 1.09) | -2.19% (-2.12%, -2.27%) |
| From E to F | 1.1115 (1.09, 1.13) | +5.39% (+3.84%, +6.95%) |
| From F to G | 0.9773 (0.96, 0.995) | -12.08% (-11.9%, -12.3%) |
| Beyond G | 1.0660 (1.05, 1.08) | +9.07% (+6.85%, +11.18%) |
| Non-Delta region, Arkansas |  |  |
| Before policy A | 2.7896 (2.36, 3.27) |  |
| From A to B | 1.0412 (0.873, 1.23) | -62.67% (-62.4%, -63.0%) |
| From B to C | 0.9309 (0.813, 1.059) | -10.81% (-26.9%, +8.35%) |
| From C to D | 1.0589 (0.979, 1.143) | +13.64% (-0.835%, +29.01%) |
| From D to E | 1.2119 (1.18, 1.24) | +14.29% (+5.14%, +23.68%) |
| From E to F | 1.0493 (1.03, 1.07) | -13.44% (-12.5%, -14.3%) |
| From F to G | 0.9342 (0.916, 0.953) | -10.97% (-10.6%, -11.3%) |
| Beyond G | 1.0701 (1.06, 1.08) | +14.51% (+12.3%, +16.7%) |

**Supplementary Table 4.**
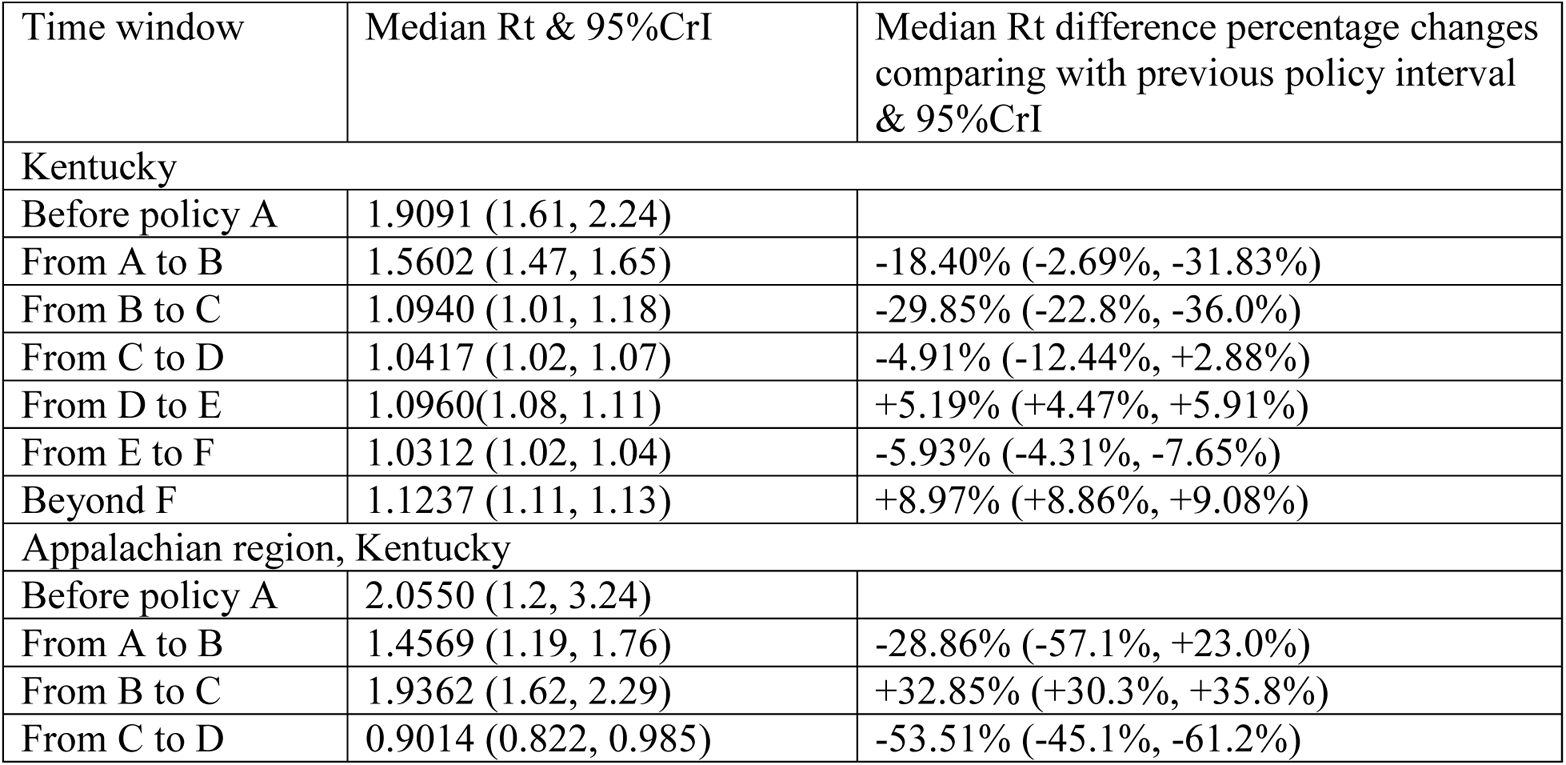
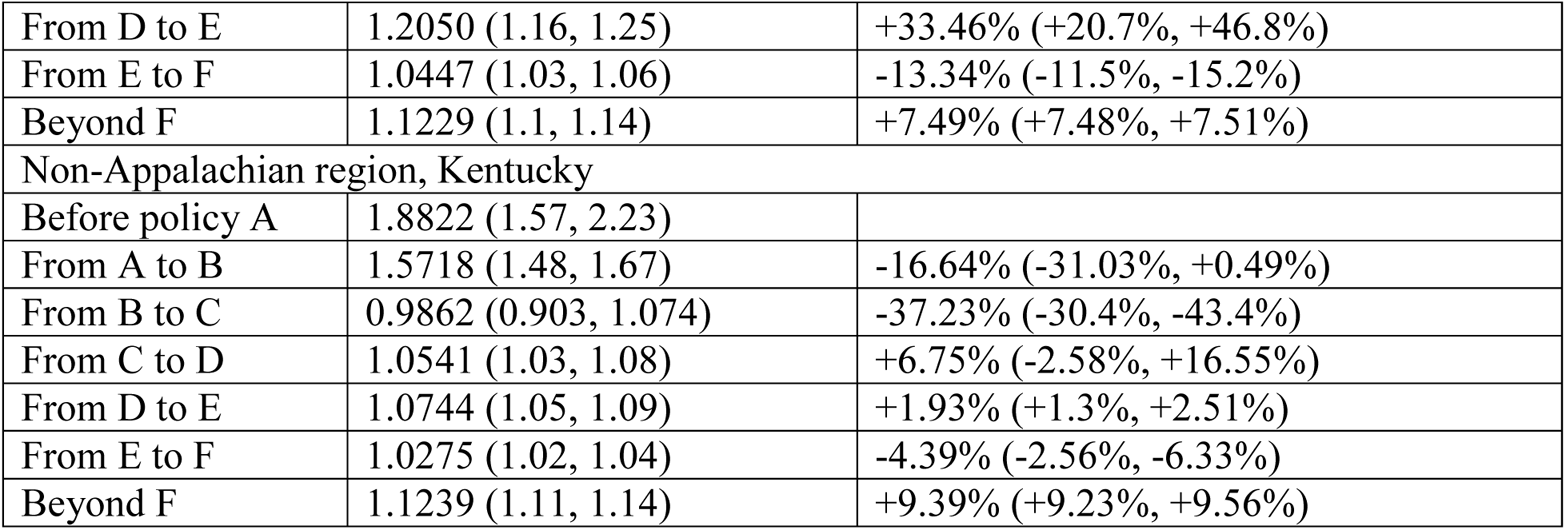
Time-varying reproduction number estimated in Kentucky using non-overlapping time windows (median and 95% credible interval) and its change between each time window (median and 95% credible interval)

**Supplementary Table 5.**
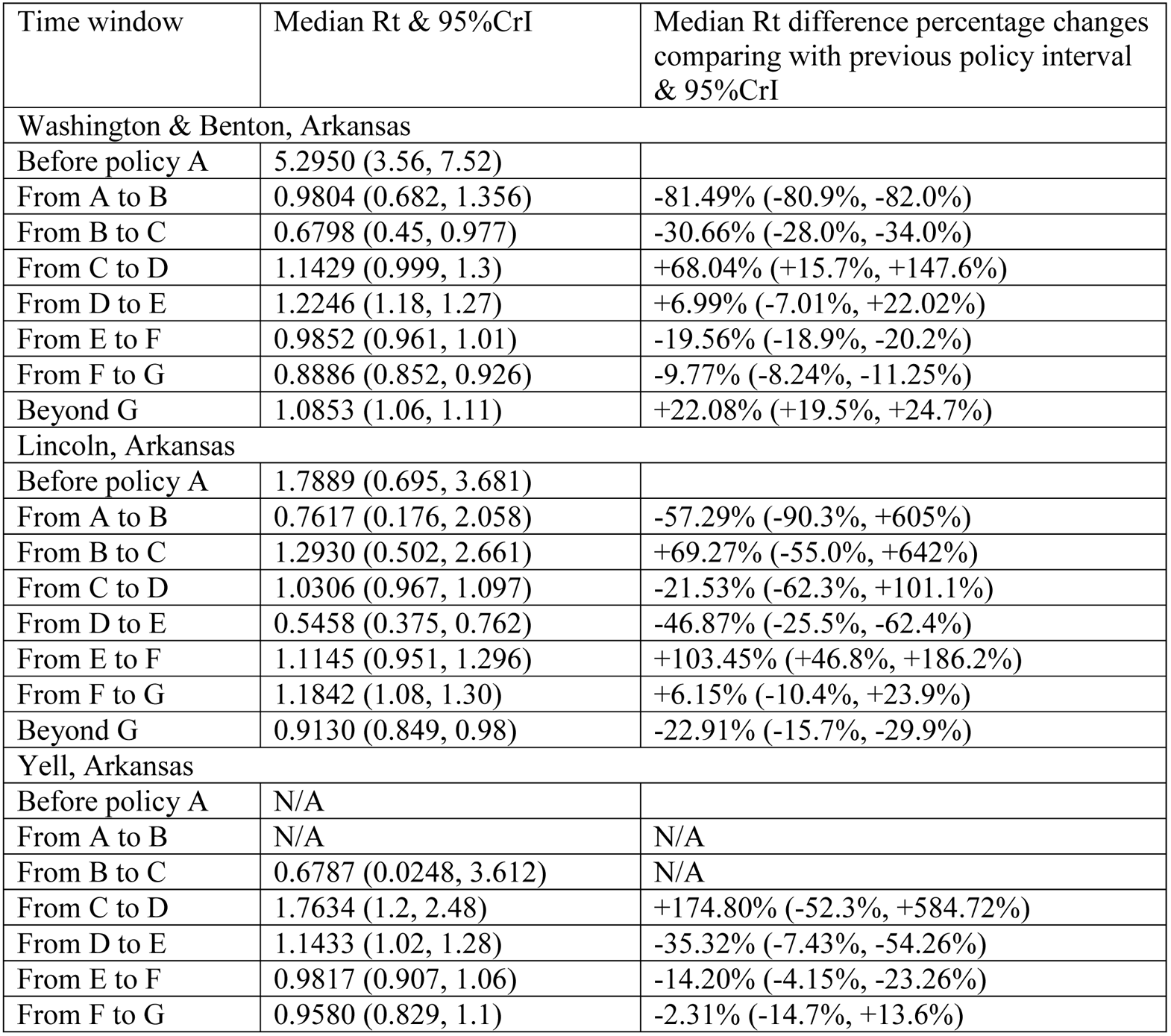
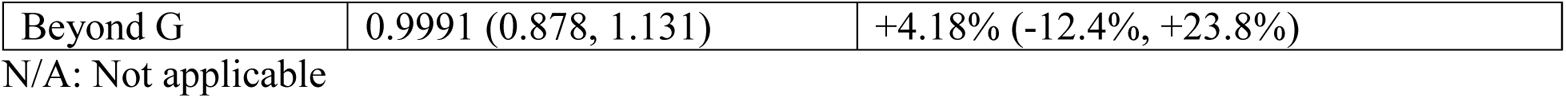
Time-varying reproduction number in four Arkansas Counties estimated using non-overlapping time windows (median and 95% credible interval) and its change between each time window (median and 95% credible interval)

| Time window | Median Rt & 95%CrI | Median Rt difference percentage changes comparing with previous policy interval & 95%CrI |
| --- | --- | --- |
| Washington & Benton, Arkansas |  |  |
| Before policy A | 5.2950 (3.56, 7.52) |  |
| From A to B | 0.9804 (0.682, 1.356) | -81.49% (-80.9%, -82.0%) |
| From B to C | 0.6798 (0.45, 0.977) | -30.66% (-28.0%, -34.0%) |
| From C to D | 1.1429 (0.999, 1.3) | +68.04% (+15.7%, +147.6%) |
| From D to E | 1.2246 (1.18, 1.27) | +6.99% (-7.01%, +22.02%) |
| From E to F | 0.9852 (0.961, 1.01) | -19.56% (-18.9%, -20.2%) |
| From F to G | 0.8886 (0.852, 0.926) | -9.77% (-8.24%, -11.25%) |
| Beyond G | 1.0853 (1.06, 1.11) | +22.08% (+19.5%, +24.7%) |
| Lincoln, Arkansas |  |  |
| Before policy A | 1.7889 (0.695, 3.681) |  |
| From A to B | 0.7617 (0.176, 2.058) | -57.29% (-90.3%, +605%) |
| From B to C | 1.2930 (0.502, 2.661) | +69.27% (-55.0%, +642%) |
| From C to D | 1.0306 (0.967, 1.097) | -21.53% (-62.3%, +101.1%) |
| From D to E | 0.5458 (0.375, 0.762) | -46.87% (-25.5%, -62.4%) |
| From E to F | 1.1145 (0.951, 1.296) | +103.45% (+46.8%, +186.2%) |
| From F to G | 1.1842 (1.08, 1.30) | +6.15% (-10.4%, +23.9%) |
| Beyond G | 0.9130 (0.849, 0.98) | -22.91% (-15.7%, -29.9%) |
| Yell, Arkansas |  |  |
| Before policy A | N/A |  |
| From A to B | N/A | N/A |
| From B to C | 0.6787 (0.0248, 3.612) | N/A |
| From C to D | 1.7634 (1.2, 2.48) | +174.80% (-52.3%, +584.72%) |
| From D to E | 1.1433 (1.02, 1.28) | -35.32% (-7.43%, -54.26%) |
| From E to F | 0.9817 (0.907, 1.06) | -14.20% (-4.15%, -23.26%) |
| From F to G | 0.9580 (0.829, 1.1) | -2.31% (-14.7%, +13.6%) |
| Beyond G | 0.9991 (0.878, 1.131) | +4.18% (-12.4%, +23.8%) |

N/A: Not applicable

**Supplementary Table 6.** Time-varying reproduction number in four Kentucky Counties estimated using non-overlapping time windows (median and 95% credible interval) and its change between each time window (median and 95% credible interval)

| Time window | Median Rt & 95%CrI | Median Rt difference percentage changes comparing with previous policy interval & 95%CrI |
| --- | --- | --- |
| Jefferson & Shelby, Kentucky |  |  |
| Before policy A | 1.8504 (1.33, 2.49) |  |
| From A to B | 1.5963 (1.44, 1.76) | -13.83% (-36.7%, +16.7%) |
| From B to C | 0.6823 (0.563, 0.817) | -57.09% (-48.1%, -64.5%) |
| From C to D | 1.0938 (1.04, 1.15) | +59.92% (+32.0%, +95.6%) |
| From D to E | 1.0184 (0.983, 1.055) | -6.91% (-5.9%, -7.93%) |
| From E to F | 1.0371 (1.02, 1.05) | +1.79% (-0.08%, +3.64%) |
| Beyond F | 1.1013 (1.08, 1.12) | +6.21% (+5.8%, +6.63%) |
| Elliott, Kentucky |  |  |
| Before policy A | N/A |  |
| From A to B | N/A | N/A |
| From B to C | N/A | N/A |
| From C to D | N/A | N/A |
| From D to E | 3.4657 (0.127, 18.444) | N/A |
| From E to F | 1.0727 (0.791, 1.414) | -67.08% (-94.3%, +723%) |
| Beyond F | 1.1748 (1.05, 1.3) | +9.23% (-18.1%, +44.6%) |
| Warren, Kentucky |  |  |
| Before policy A | 1.5157 (0.589, 3.119) |  |
| From A to B | 1.9666 (1.31, 2.81) | +29.33% (-42.9%, +253.7%) |
| From B to C | 0.9044 (0.581, 1.448) | -54.23% (-14.9%, -76.2%) |
| From C to D | 1.1545 (1.08, 1.23) | +27.06% (-21.4%, +118.9%) |
| From D to E | 0.9787 (0.918, 1.042) | -15.23% (-15.0%, -15.5%) |
| From E to F | 1.003 (0.965, 1.041) | +2.37% (-5.07%, +10.0%) |
| Beyond F | 1.1188 (1.06, 1.18) | +11.59% (+7.45%, +16.83%) |

N/A: Not applicable

**Supplementary Table 7.** The slope (and 95% Confidence Intervals) of the regression line between per capita cumulative case count and population size, by region, Arkansas and Kentucky, on May 7, July 7, September 7 and November 7, 2020.

|  | May 7 | July 7 | September 7 | November 7 |
| --- | --- | --- | --- | --- |
| Appalachian | 0.05 (-0.43, 0.52) | 0.39 (0.10, 0.68) | 0.18 (-0.02, 0.39) | -0.01 (-0.14, 0.12) |
| Non-Appalachian | 0.29 (0.07, 0.52) | 0.24 (0.07, 0.41) | 0.13 (0.02, 0.23) | 0.10 (0.02, 0.18) |
| Delta | 0.24 (-0.25, 0.73) | 0.13 (-0.29, 0.56) | 0.02 (-0.28, 0.32) | 0.02 (-0.18, 0.22) |
| Non-Delta | 0.04 (-0.32, 0.40) | 0.28 (-0.08, 0.64) | 0.09 (-0.10, 0.29) | 0.05 (-0.09, 0.18) |

